# Neutralizing Antibody to Omicron BA.1, BA.2 and BA.5 in COVID-19 Patients

**DOI:** 10.1101/2022.08.21.22278552

**Authors:** Susanne L. Linderman, Lilin Lai, Estefany L. Bocangel Gamarra, Nicholas M. Mohr, Kevin W. Gibbs, Jay S. Steingrub, Matthew C. Exline, Nathan I. Shapiro, Anne E. Frosch, Nida Qadir, Srilatha Edupuganti, Diya Surie, Mark W. Tenforde, Meredith E. Davis-Gardner, James D. Chappell, Max S Y Lau, M. Juliana McElrath, Adam S. Lauring, Mehul S. Suthar, Manish M. Patel, Wesley H. Self, Rafi Ahmed

## Abstract

Neutralizing antibody plays a key role in protective immunity against COVID-19. As increasingly distinct variants circulate, debate continues regarding the value of adding novel variants to SARS-CoV-2 vaccines. In this study, we have analyzed live virus neutralization titers against WA1, Delta, BA.1, BA.2, and BA.5 in 187 hospitalized patients infected with Delta or Omicron strains. This information will be useful in selection of the SARS-CoV-2 strains to include in an updated vaccine. Our results show that unvaccinated Delta infected patients made a highly biased neutralizing antibody response towards the infecting Delta strain with slightly lower responses against the WA1 strain, but with strikingly lower titers against BA.1, BA.2, and BA.5. Delta infected patients that had been previously vaccinated with the WA1 containing COVID vaccine made equivalent responses to WA1 and Delta strains, but still had very low neutralizing antibody responses to Omicron strains. In striking contrast, both unvaccinated and vaccinated Omicron patients exhibited a more balanced ratio of Omicron virus neutralization compared to neutralization of ancestral strains. Interestingly, Omicron patients infected with BA.1 or BA.2 had detectable neutralizing antibody titers to BA.5, but these titers were lower than neutralization titers to BA.1 and BA.2. Taken together, these results suggest that inclusion of the Omicron BA.5 strain in a SARS-CoV-2 vaccine would be beneficial in protection against the widely circulating BA.5 variant.

**Disclaimer:** The findings and conclusions in this report are those of the authors and do not necessarily represent the official position of the Centers for Disease Control and Prevention.

---

Neutralizing antibody plays a key role in protective immunity against SARS-CoV-2 (1). This is evident from the large number of Omicron infections in vaccinated and convalescent patients since antibodies induced by vaccination or infection by the ancestral WA1 strain do not neutralize the Omicron variants efficiently (2, 3). This has led to the recommendation by the FDA for including the Omicron variant in COVID-19 vaccines (4). However, issues have been raised about the value of adding Omicron to the vaccine based on data showing only modest differences between antibody responses after booster immunization with Omicron versus WA1 (5-7). Questions also remain about which Omicron variant(s) should be used and whether the WA1 strain should still be kept in the updated vaccine.

Having more detailed information about the neutralizing antibody profiles after infection with different SARS-CoV-2 variants will be helpful in making these critical decisions about vaccine selection. Here we report live virus neutralization titers against WA1, Delta, BA.1, BA.2 and BA.5 variants in serum samples collected from hospitalized patients infected with Delta or Omicron strains. Blood samples were collected from 187 patients hospitalized with acute COVID-19 between July 2021 – March 2022 at 8 U.S. hospitals (Supplemental Methods). The majority (69%) of these patients were sequence confirmed for Delta or Omicron infection and the remaining were classified according to calendar period of circulation (Supplemental Table 1). Patients were either unvaccinated (n=80) or vaccinated with COVID-19 mRNA vaccine (n=100) or adenovirus vector vaccine (n=7) before infection.

Unvaccinated Delta infected patients made a highly biased neutralizing antibody response towards the infecting Delta strain with lower titers against WA1 (6-fold) and strikingly lower titers against BA.1 (60-fold) and BA.2 (22-fold) (Figure 1A). In vaccinated Delta patients the neutralization titers were similar between Delta and WA1 most likely reflecting the recruitment of memory B cells induced by the WA1 vaccine (Figure 1B), but once again, neutralizing activity against BA.1 and BA.2 was much lower (17-fold and 9-fold respectively, Figure 1B). Thus, both unvaccinated and vaccinated Delta infected patients made significantly lower neutralizing antibody responses to Omicron (as determined by Wilcoxon rank sums test). A strikingly different pattern was seen in Omicron infected patients irrespective of their vaccination history with a more favorable neutralizing antibody response to BA.1 and BA.2. While there was some variation between individual samples, there was a clear trend for a broader and more balanced antibody response with similar neutralization titers to Delta, BA.1, and BA.2 (Figures 1C, 1D). Additionally, while neutralization titers to the vaccine strain WA1 were moderately higher (3-fold) than BA.1 and BA.2 neutralizing titers in vaccinated Omicron patients at this time, the ratio of BA.1 to WA1 neutralizing titers was significantly higher in Omicron patients compared to Delta patients in both unvaccinated (19-fold) and vaccinated (10-fold) patients (Figure 1E, 1F). This analysis of the ratio between neutralizing antibody to omicron versus WA1 within a given individual nicely documents that omicron infection favors omicron specific antibody responses.

**Figure 1:**
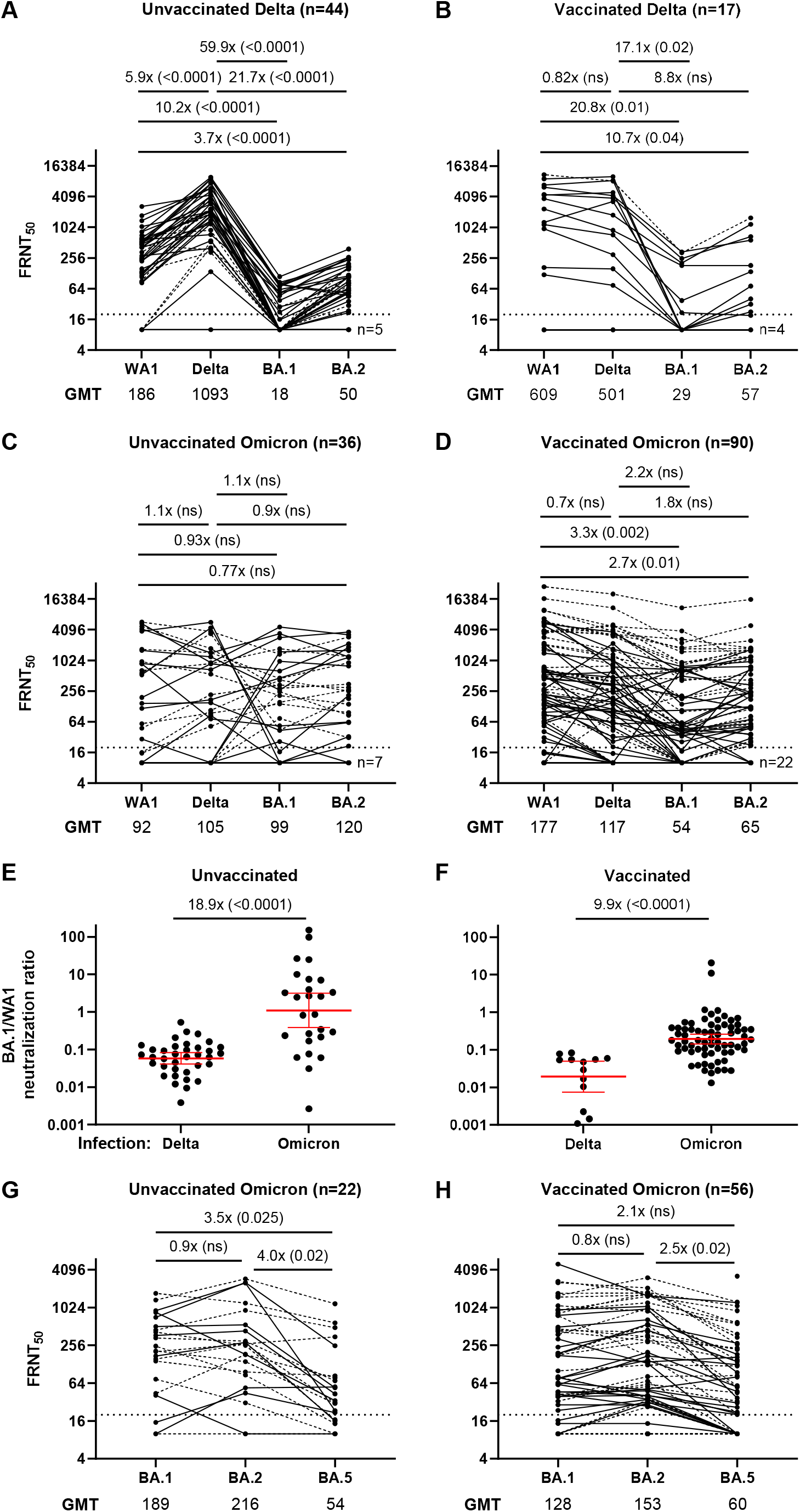
Live virus neutralizing antibody titers in hospitalized patients infected with SARS-CoV-2 Delta or Omicron variants. In vitro neutralization titers (FRNT50 or focus reduction neutralization test) against live WA1, Delta, BA.1, or BA.2 variant in unvaccinated (A) and vaccinated (B) hospitalized patients infected with a Delta strain. Geometric mean titers (GMTS) are listed below each panel and the ratio of GMTs is shown above the panel. Solid connecting lines indicate samples from patients with sequence confirmed Delta infection, while dashed lines indicate samples that were not sequence confirmed but collected while Delta was the dominant circulating strain. Limit of detection of 20 is indicated by a dotted line. In vitro neutralization titers against live WA1, Delta, BA.1, or BA.2 variant in unvaccinated (C) and vaccinated (D) hospitalized patients infected with an Omicron strain. Solid connecting lines indicate samples from patients with sequence confirmed Omicron infection, while dashed lines indicate samples that were not sequence confirmed but collected while Omicron was the dominant circulating strain. The ratio of BA.1 to WA1 neutralizing titers in unvaccinated (E) or vaccinated (F) individuals infected with Delta or Omicron strains. Geometric mean and 95% CI are indicated in red. For a subset of Omicron patients who were unvaccinated (G) or vaccinated (H), BA5 neutralization was also assessed. All p values were calculated by Wilcoxon rank sum test with continuity correction.

Given the importance of the currently dominant BA.5 strain we then tested neutralization against BA.5. Note that our samples were collected prior to dominance of BA.5 and most of our patients were infected with BA.1 (Supplemental Table 1). In the subset of Omicron infected samples that we analyzed, there was detectable neutralization of BA.5 but it was lower than for BA.1. and BA.2 in both vaccinated and unvaccinated patients (Figure 1G, 1H, Supplemental Figure 1). This reduced neutralization activity against BA.5 in patients infected with BA.1/BA.2 supports the inclusion of BA.5 in the vaccine.

In summary, our results show that Omicron infection of unvaccinated or vaccinated patients induces a more proportional and balanced neutralizing antibody response to Omicron variants supporting the recent decision to include an Omicron lineage strain in a SARS-CoV-2 vaccine. However, it remains to be seen how these findings from infection will translate to vaccination. Our results are from hospitalized patients who most likely had high levels of infected cells and antigen that would have efficiently induced a primary response to Omicron. It is unlikely that a single immunization with an Omicron vaccine would do the same and two shots may be needed to prime sufficient naïve B cells to give the desired antibody response. Also, it needs to be considered whether keeping the ancestral WA1 strain in the updated vaccine is of value or not.

## Data Availability

All data produced in the present study are available upon reasonable request to the authors

## Supplemental Materials

### Supplemental Methods

#### Surveillance

This prospective, multicenter observational assessment was conducted by the Influenza and Other Viruses in the Acutely Ill (IVY) Network in collaboration with the US Centers for Disease Control and Prevention (CDC). The IVY Network is a 21-hospital collaborative in the US studying COVID-19. Eight hospitals participated in the serology component included in the current analysis. The current analysis included adult patients hospitalized at IVY sites during the period of July 4, 2021–March 30, 2022. Site personnel prospectively identified and enrolled hospitalized COVID-19 case-patients. Participants were adults (≥18 years old) admitted to the hospital with symptomatic COVID-19 confirmed with a positive SARS-CoV-2 reverse transcription-polymerase chain reaction (RT-PCR) test within 14 days of symptom onset. Participants had one or more of the following COVID-19-associated signs or symptoms: fever, cough, shortness of breath, loss of taste, loss of smell, use of respiratory support (high flow oxygen by nasal cannula, non-invasive ventilation, or invasive mechanical ventilation) for the acute illness, or new pulmonary findings on chest imaging indicating pneumonia. For confirmation of SARS-CoV-2 infection, nasal specimens were centrally tested at Vanderbilt University Medical Center by RT-qPCR for SARS-CoV-2 nucleocapsid 1 and nucleocapsid 2 targets using CDC designed primers and probes. Specimens positive for SARS-CoV-2 were sent to University of Michigan for viral whole genome sequencing as described below. For respiratory specimens that could not be sequenced, periods of predominant circulation for Delta, and Omicron were defined based on time windows when each variant was identified in more than 50% of cases successfully sequenced in the study— Delta period: 4 July to 25 December 2021; Omicron period: 26 December 2021 to March 30, 2022. Trained personnel at enrolling sites collected patient data on demographics, medical history, underlying health conditions, COVID-19 vaccination status, and clinical outcomes through patient or proxy interviews and medical record review. A convenience sample of patients had a serum specimen during hospitalization. These activities were reviewed by CDC, were conducted consistent with applicable federal law and CDC policy (45 C.F.R. part 46.102(l)(2), 21 C.F.R. part 56; 42 U.S.C. §241(d); 5 U.S.C. §552a; 44 U.S.C. §3501 et seq), and were determined to be public health surveillance with waiver of informed consent by institutional review boards at CDC and each enrolling site.

#### Molecular diagnosis and sequencing

Upper respiratory specimens with detection of either N1 or N2 with a cycle threshold ≤32 were shipped to the University of Michigan (Ann Arbor, Michigan) for viral whole genome sequencing using the ARTIC Network protocol (v4.1 primer set) and an Oxford Nanopore Technologies GridION instrument.

#### Viruses and cells

VeroE6-TMPRSS2 cells were generated and cultured as previously described(1). nCoV/USA_WA1/2020 (WA/1), closely resembling the original Wuhan strain was propagated from an infectious SARS-CoV-2 clone as previously described(2). icSARS-CoV-2 was passaged once to generate a working stock. The B.1.617.2, BA.1 and BA.2 variants were isolated and propagated as previously described (1, 3). The BA.5 isolate was kindly provided by Dr. Richard Webby (St Jude Children’s Research Hospital) was plaque purified and propagated once in VeroE6-TMPRSS2 cells to generate a working stock. All viruses used in this study were deep sequenced and confirmed as previously described (1).

#### Focus Reduction Neutralization Test

FRNT assays were performed as previously described (1, 4, 5). Briefly, samples were diluted at 3-fold in 8 serial dilutions using DMEM (VWR, #45000-304) in duplicates with an initial dilution of 1:10 in a total volume of 60 μl. Serially diluted samples were incubated with an equal volume of WA1/2020, B.1.617.2 (Delta), or BA.1, BA.2 or BA.5 (Omicron) with 100-200 foci per well at 37° C for 1 hour in a 96-well plate. The antibody-virus mixture was then added to VeroE6-TMPRSS2 cells and incubated at 37°C for 1 hour. Post-incubation, the antibody-virus mixture was removed and 100 µl of pre-warmed 0.85% methylcellulose overlay was added to each well. Plates were incubated at 37° C for 18 to 40 hours and the methylcellulose overlay was removed and washed six times with PBS. Cells were fixed with 2% paraformaldehyde in PBS for 30 minutes. Following fixation, plates were washed twice with PBS and permeabilization buffer (0.1% BSA, 0.1% Saponin in PBS) was added to permeabilized cells for at least 20 minutes. Cells were incubated with an anti-SARS-CoV spike primary antibody directly conjugated to Alexaflour-647 (CR3022-AF647) overnight at 4°C. Cells were then washed twice with 1x PBS and imaged on an ELISPOT reader (CTL Analyzer).

#### Quantification and Statistical Analysis

Antibody neutralization was quantified by counting the number of foci for each sample using the Viridot program (6). The neutralization titers were calculated as follows: 1 - (ratio of the mean number of foci in the presence of sera and foci at the highest dilution of respective sera sample). Each specimen was tested in duplicate. The FRNT-50 titers were interpolated using a 4-parameter nonlinear regression in GraphPad Prism 9.2.0. Samples that do not neutralize at the limit of detection at 50% are plotted at 20 and was used for geometric mean and fold-change calculations. The Wilcoxon rank sum test was used for testing differences between groups. Whenever applicable, corrections for multiple testing were performed using the p-value adjustment method by Holm (7).

**Supplemental Figure 1.**
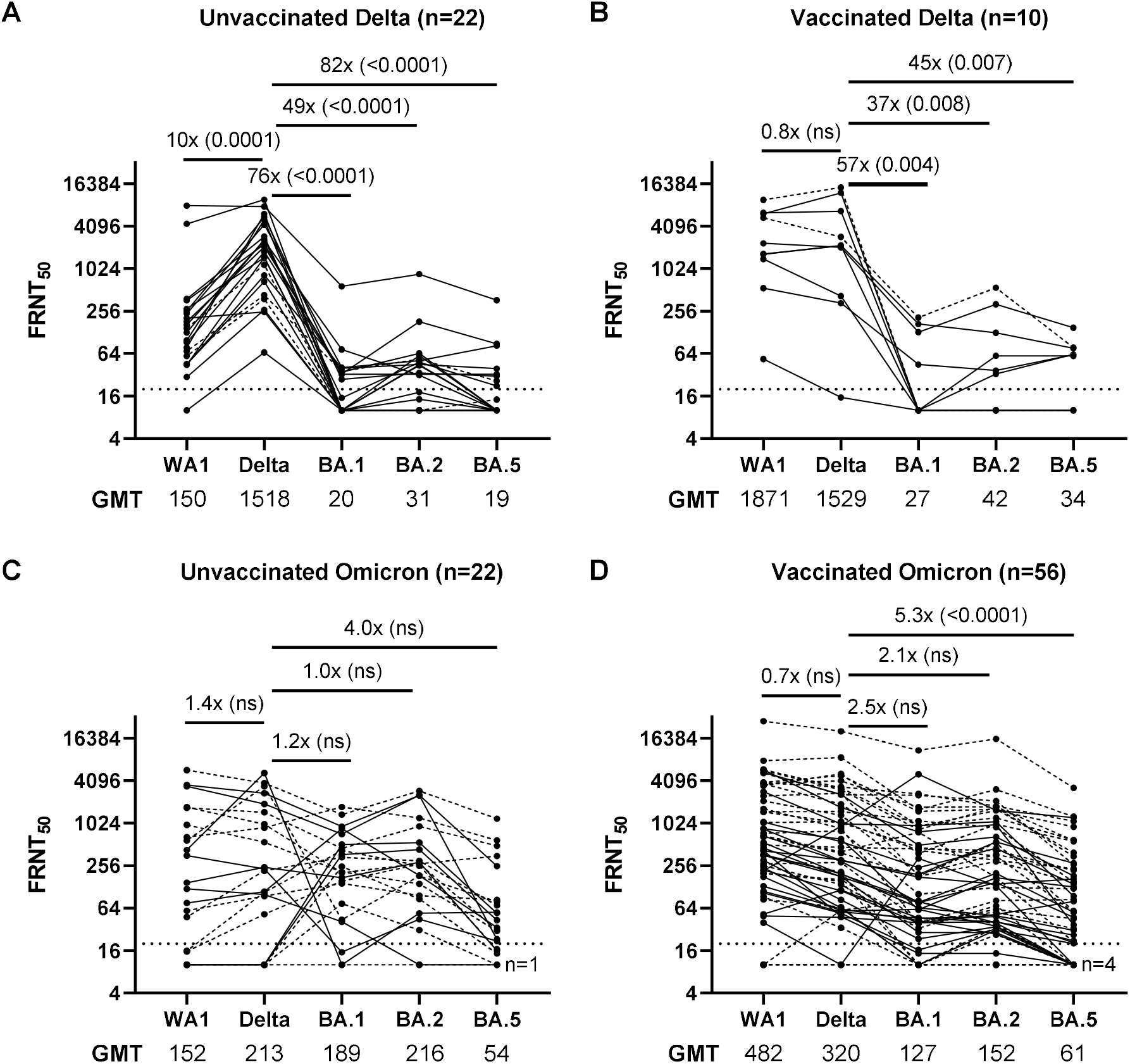
Live virus neutralizing antibody titers in hospitalized patients infected with SARS-CoV-2 Delta or Omicron variants. I*n vitro* neutralization titers (FRNT50 or focus reduction neutralization test) against live WA1, Delta, BA.1, BA.2, or BA.5 virus in unvaccinated (A) and vaccinated (B) hospitalized patients infected with a Delta strain. Geometric mean titers (GMT) are listed below each panel. The limit of detection of 20 is indicated by a dotted line. *In Vitro* neutralization titers against live WA1, Delta, BA.1, BA.2, or BA.5 virus in unvaccinated (C) and vaccinated (D) hospitalized patients infected with an Omicron strain. Solid connecting lines indicate samples from patients with sequence confirmed infection, while dashed lines indicate samples that were not sequence confirmed but assigned to a group due to the prevalence of contemporary strains.

**Supplemental Table 1.**
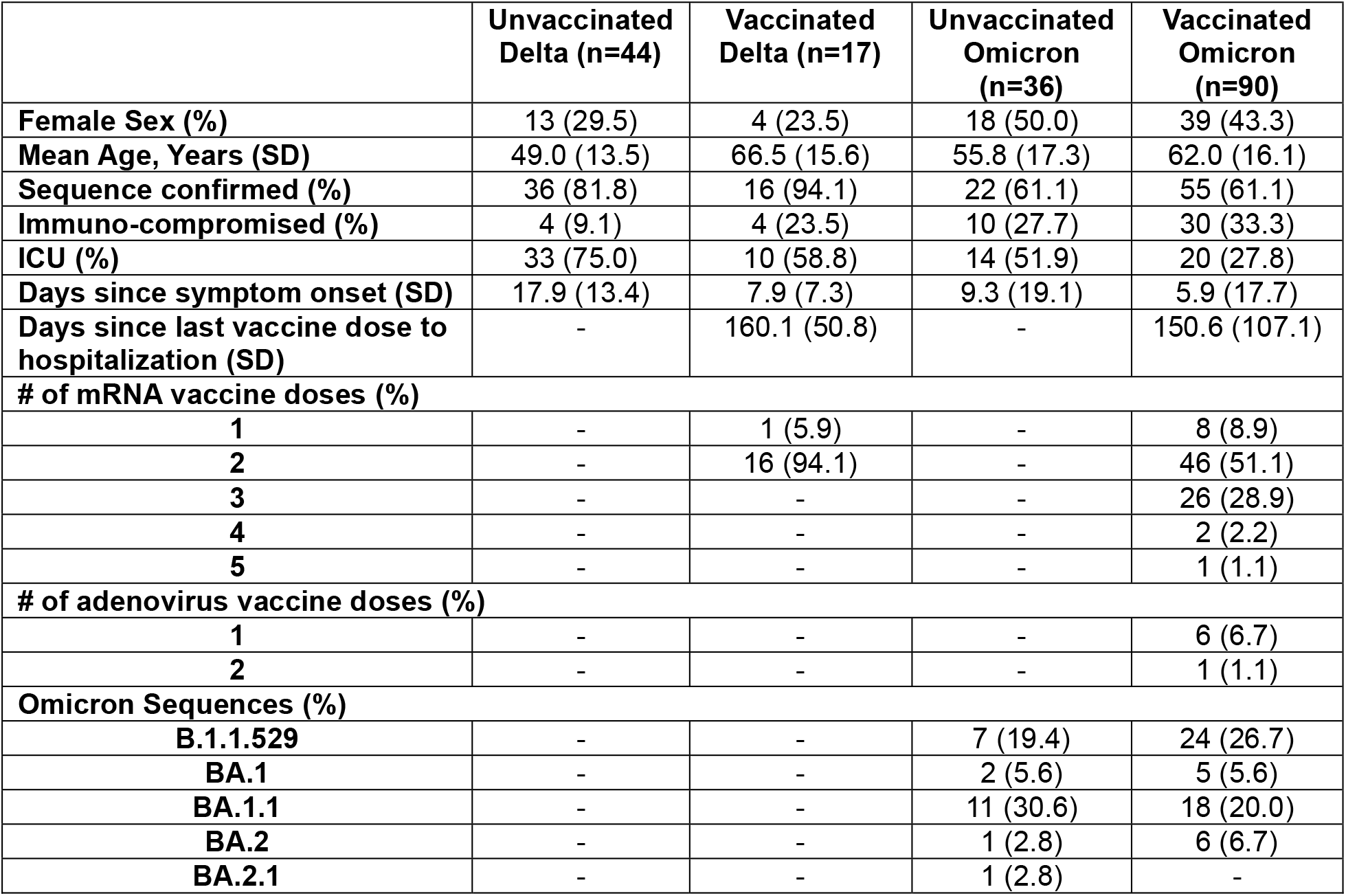
Demographic and clinical cohort characteristics of adult inpatients hospitalized with COVID-19 who provided serum for analysis.

